# Synthetic Spatiotemporal Covid-19 Data Replicates Generated via the USPopulationSampler R Package

**DOI:** 10.64898/2026.08.20.26360919

**Authors:** Rithy Techavoan Yean, Jasen Zhang, Andrew J. Holbrook

**Affiliations:** University of California, Los Angeles, Department of Biostatistics, Fielding School of Public Health, USA

## Abstract

Given that a disease case is known to have occurred in a specific county or state, can one reconstruct a precise geographic observation for that case? The answer is that one cannot. We instead sample multiple high-probability locations for that case under the assumption that cases are more likely to occur in places where there are more people. To do this, we develop a novel and user-friendly `R` package called `USPopulationSampler` that randomly samples geospatial locations within census block groups (BG; the smallest geographic unit for which population counts are available) within target counties, states, or across the entirety of the U.S., randomly selected according to population counts using Census Bureau reference data. We use the package to generate 28 realistic high-probability data replicates of 100M+ synthetic spatiotemporal Covid-19 cases observed between the dates of January 21st, 2020 to March 23rd, 2023 and openly publish these data replicates on `Zenodo` for easy access. Given the large scale nature of the data, the `USPopulationSampler` package also provides tools for fast download of these datasets and functions to generate further replicates at scale using multi-core parallelization.

## Background and Summary

Spatiotemporal point data provide both the location and the time of occurrence of discrete events of interest within some designated spatial region^1^. There is extensive literature on the statistical modeling of spatiotemporal event data, with spatiotemporal point processes providing the fundamental framework for describing, estimating, and predicting the occurrence of events across space and time^1,2^. Applications of spatiotemporal point process models span diverse fields, with self-exciting Hawkes processes modeling infectious disease transmission and incidence, epidemic-type aftershock sequence models characterizing spatial and temporal clustering of earthquakes, and Poisson hurdle processes predicting wildfire ignitions^3–5^. In parallel with advances in statistical modeling of spatiotemporal event data, there is also a growing body of literature applying deep learning and other AI methods on spatiotemporal event data across domains in criminology, seismography, and human mobility ^6–9^. Precise event locations and timestamps have thus become crucially important in enabling the learning of complex spatial and temporal patterns arising from individual event phenomena.

Unfortunately, many of this existing and developing technology cannot be used to improve our knowledge of forecasting epidemics’ spatial dynamics because of the often spatially coarse nature of epidemiological data. Disease events are reported as counts within administrative areas rather than at their individual exact locations causing spatial aggregation of surveillance data^10^. For instance, common public data sources for coronavirus disease 2019 (Covid-19) data often aggregates cases per state, county, or city by health agencies and state departments^11–13^. While Covid-19 in the U.S. was monitored in near real time by *The New York Times (NYT)* from January 21, 2020 to March 24, 2023, reported cases and deaths were provided as cumulative counts aggregated to the county level^12^. However, cumulative counts data is unlikely to differentiate regional population variation especially for disease surveillance^14^. Given only that a disease was reported within a particular administrative region, such as a county or state, the exact location of that infection cannot be recovered because the original spatial information has been lost through aggregation or the data collection process. Nevertheless, it is instead possible to generate statistically plausible realizations of locations by incorporating external information about the underlying population distribution. The proposed framework is to assume that more cases are likely to appear in places where there are more people. Densely populated areas are positively linked to higher number of observed Covid-19 cases among other factors that contribute to the contagion spread of the disease^15^. Epidemiological individual-level data is therefore much needed during pandemics to best anticipate the spread of infection by providing both location and time data to allow descriptive mapping of occurrences through time while enabling estimations of epidemiological parameters using mathematical models^16^.

Even though 95% of U.S. hospitals have the electronic health record systems, the U.S. lacks a national public reporting system architecture that is free of inconsistencies and data discrepancies caused by decentralized data aggregation and collection^17^. To accommodate a change in reporting, data collecting, and disease surveillance at the national level, moving from aggregated counts to individual-level would be very challenging due to limited resources and privacy considerations. Synthetic data have become much more popular for simulation studies and is used in several health care domains^18^. This motivates the creation of the `USPopulationSampler` package in R which provides a method to generate synthetic spatiotemporal disease data. Given how large scale these synthetic data replicates can be, tools are provided in `USPopulationSampler` that allow users to enable multi-core parallelization to generate further replicates of their own interest^19^.

In particular, `USPopulationSampler` samples multiple high-probability locations under the assumption that cases are more likely to occur in places where there are many people^19^. The package samples locations uniformly in census block groups that are randomly selected according to population counts of block groups across the United States of America by using the 2020 Decennial U.S. Census Block Maps data as a reference dataset^20^. Existing spatial analysis packages like the simple functions (`sf`) package provide efficient tools for representing, manipulating, and sampling from spatial vector data in the `R` environment^21,22^. Namely, the `st_sample()` function enables random point generation within arbitrary polygon geometries ensuring locations are guaranteed to lie inside the spatial boundaries of an object^21^. However, while the `sf` package provides the geometric framework for sampling within polygons, it does not allocate samples according to population sizes or consider demographic information. The `USPopulationSampler` fills that need by integrating the sf package directly into its sampling architecture enabling the user to allocate points across the 50 states within the U.S. as well as the District of Columbia and the territory of Puerto Rico at the block group granularity. Block groups are subdivisions of a census county typically with population between 600 to 3,000 and are the smallest geographic entity that maps distinct neighborhoods for which the U.S. Census Bureau publishes and tabulates its geospatial data^23^. Therefore, the `USPopulationSampler` distributes spatial observations that closely reflect the spatial distribution of the population while preserving the heterogeneity of residential areas in the US.

## Data Overview

The spatial distribution of synthetic locations differs among replicates because the locations are generated by random sampling census block groups according to population counts. The synthetic datasets cover 3,210 U.S. counties with reported Covid-19 cases between January 21st, 2020 to March 23rd, 2023.

**Figure 1.**
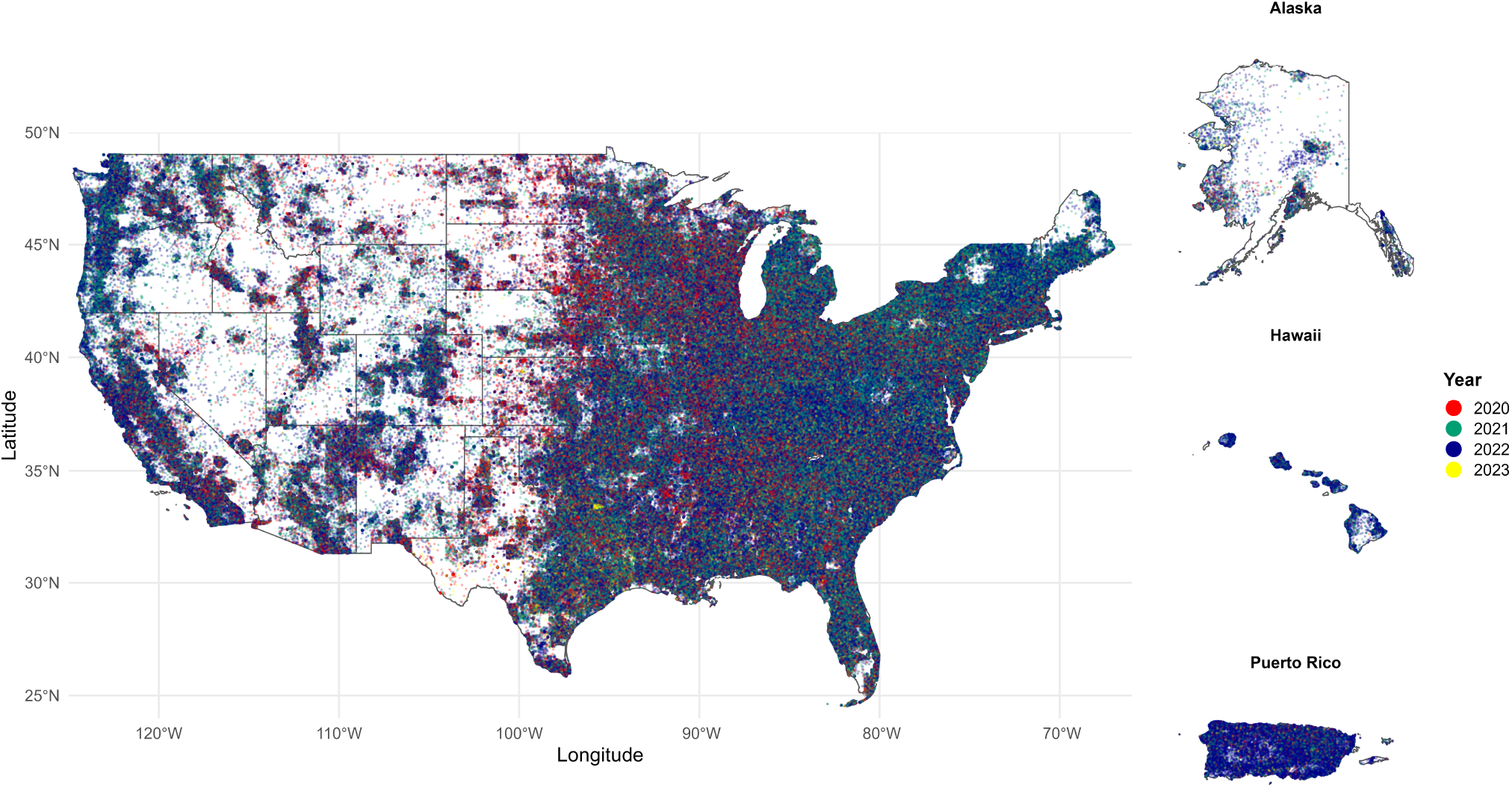
Synthetic spatiotemporal Covid-19 locations from data replicate No. 18 with (10M+ of 100M+) points plotted

## Methods

We explore the mathematical logic behind the sampling framework used in the `US_pop_sampler()` function provided in the `USPopulationSampler` package. Built-in functions `load_bg_data()` and `load_covid_data()` load the reference datasets for usage. For interests concerning the up-to-date reconstruction of these reference datasets, please refer to the chapters outlined in the README^24^.

### Simulating Synthetic Spatiotemporal Covid-19 Data Replicates

Once the reference datasets have been prepared, we now discuss the generation of the synthetic spatiotemporal Covid-19 data replicates using the functionalities built within the USPopulationSampler package. The three functions of interest are the `US_pop_sampler()` function, the `assign_dates_to_pts()` function, and the `open_synthetic_data()` function. The mathematical frameworks for `US_pop_sampler()` function and the `assign_dates_to_pts()` are discussed below:

#### Sampling Mathematical Framework

The following mathematical framework describes the generation of the synthetic COVID-19 spatiotemporal data replicates using the exact Federal Information Processing Series (FIPS) code count sampling and temporal assignment procedures implemented in the package. FIPS codes are standardized numbers published by U.S. Census Bureau and developed by the National Institute of Standards and Technology that uniquely identify geographic areas from states to census block groups^25,26^. Let B represent the set of all census block groups in the 2020 US Decennial Block Maps Reference dataset represented as *B* = *{*1, 2, …, *m}* ^20, 23^. For each block group *i*, let *P*_*i*_ denote its population count. Only block groups satisfying the condition *P*_*i*_ *>* 0 are eligible for sampling. The total population across all valid block groups is thus denoted by 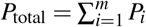.

If a user requests *N* synthetic observations, then the probability of selecting block group *i*, is *p*_*i*_ = *P*_*i*_*/P*_total_. The vector of allocated counts (*C*_1_,*C*_2_, …,*C*_*m*_) is sampled from a multinomial distribution (*C*_1_,*C*_2_, …,*C*_*m*_) *∼* Multinomial(*N, p*_1_, *p*_2_, …, *p*_*m*_). This guarantees that the sum of all allocated counts is equal to *N* which is represented as 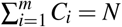. The expected number of sampled observations within block group *i* is *E*[*C*_*i*_] = *Np*_*i*_. Consequently, larger population block groups receive proportionally more synthetic observations than sparsely populated areas as their probability of being selected is higher. Once the block group counts have been allocated, the synthetic spatial locations are generated independently within each block group’s polygon region. A polygon region is simply the region of that county’s block group which consists of several spatial coordinates that identify the shape of that place.

Let *G*_*i*_ denote the geographic polygon associated with census block group *i*. For each allocated observation *j* within block group *i, X*_*ij*_ = (*x*_*ij*_, *y*_*ij*_) is sampled uniformly within the polygon interior *X*_*ij*_ *∼* Uniform(*G*_*i*_). Locations are therefore independently sampled as follows: 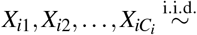 Uniform(*G* ).

Some reported county FIPS codes may not be present within the census block group dataset. Let *M* denote the the set of missing county FIPS codes. For each missing county *f ∈ M*, let *N*_*f*_ denote the number of observations associated with that county. Rather than discarding these observations, the algorithm performs a hierarchical reconciliation procedure. For each missing county *f*, let *S* _*f*_ denote the state identified by the first 2 digits of the county FIPS code *f* . If *S* _*f*_ corresponds to a valid state in the reference dataset, observations are redistributed only among counties within that state. For each county *c ∈ S*_*f*_ the county population weight is 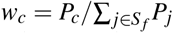 where *P*_*c*_ denotes the total county population of county *c* and *P*_*j*_ denote the total population of county *j* where j indexes every county belonging to *S*_*f*_ . The missing observations are then allocated according to 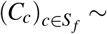 *Multinomial* 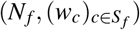, where the reconciliation is unbiased *E*[*C*_*c*_] = *N*_*f*_ *w*_*c*_ under the assumption that expected disease observations in a location are proportional to the population size of that location. The resulting allocation also satisfies 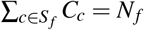 by redistributing all *N*_*f*_ observations among all valid counties without creating or removing observations. This allocation step is necessary for the FIPS codes `02261`, `02997`, and `02998` to be redistributed into valid counties in Alaska. Moreover, this allocation step allows for FIPS code `48999` to be redistributed into valid counties in Texas which preserves the total state geographic distribution of Covid-19 cases. Mathematically, this is represented as 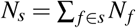 where *N*_*s*_ is the observed number of cases in state *s* and let 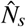 be the synthetic total. Hence, 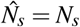 for every state whose FIPS codes exist in the census reference dataset. In the case that the specified FIPS code does not correspond to any state in the reference dataset, observations are redistributed nationally. FIPS data often uses FIPS code `99999` when FIPS code designations are missing or unavailable. In our generation of the synthetic data replicates, FIPS code `99999` was not present but the framework can be applied for such cases. For each state *s*, let 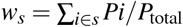 where the numerator is the total population of states *s*, and the denominator is the total population across all states. The population is first allocated among states (*C*_*s*_)_*s*_ *∼ Multinomial*(*N*_*f*_, (*w*_*s*_)_*s*_) subject to ∑_*s*_ *C*_*s*_ = *N*_*f*_ . Within each selected state, observations are subsequently redistributed among counties using county population weights 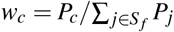.

Once the synthetic locations have been generated, dates may be assigned to them. Let *Y*_*f*_ (*t*) denote the observed number of reported Covid-19 cases in county *f* on date *t*. For county *f, N*_*f*_ = ∑_*t*_ *Y*_*f*_ (*t*) is the total number of reported cases which is equal to the number of synthetic observations generated for county *f* . This ensures that the total sample size requested is always preserved after reconciliation 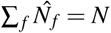. After spatial synthetic locations have been generated, the synthetic observations are randomly permuted to remove any ordering introduced during the sampling process. Let *π*_*f*_ denote a random permutation of the *N*_*f*_ synthetic observations in county *f* . The observed daily case counts are then expanded into a deterministic sequence of dates: *D*_*f*_ = {*t*_1_, …, *t*_1_, *t*_2_, …, *t*_2_, *t*_*T*_, … *t*_*T*_} where each calendar date appears exactly as many times as the reported number of cases on that date represented as |*D*_*f*_ | = ∑_*t*_ *Y*_*f*_ (*t*) = *N*_*f*_ . This is a bijection that ensures that every synthetic observation receives exactly one date. The dates are assigned sequentially to the randomly permuted observations *T*_*πf* (*k*)_ = *D*_*f*_ (*k*), *k* = 1, 2, …, *N*_*f*_ where *T*_*πf* (*k*)_ denotes the assigned date of the *kth* synthetic observation after permutation.

Consequently, 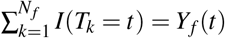 where *I*(.) is the indicator function. Therefore, for every county *f, Ŷ*_*f*_ (*t*) = *Y*_*f*_ (*t*). The only exception occurs for observations originating from county FIPS codes that cannot be spatially sampled because no corresponding census block groups exist. During the spatial reconciliation step, these observations are reassigned to valid recipient counties. Once reassigned, they inherit the expanded reporting-date sequence of the recipient county. If the number of reassigned observations does not exceed the number of available observed dates in the recipient county, dates are sampled without replacement so that each observed event is used at most once.

Suppose observations from an invalid county FIPS code *f ∈ M* are reassigned to a valid recipient county *g*. Let 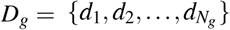 denote the expanded event-level reporting date sequence for county *g*, where |*D*_*g*_| = *N*_*g*_ = ∑_*t*_ *Y*_*g*_(*t*). Let *C*_*g*_ denote the number of synthetic observations requiring dates after reconciliation. If *C*_*g*_ *≤* |*D*_*g*_|, reporting dates are sampled uniformly without replacement from *D*_*g*_ ensuring that each observed event is used at most once. Otherwise, reporting dates are sampled with replacement *D*_*g*_. Consequently, the reconciled counties inherit the empirical temporal distribution of their recipient county while ensuring that every synthetic observation receives a valid reporting date. It should be noted that although the synthetic Covid-19 observations satisfy *N*_*f*_ = *D* for directly sampled counties, the `assign_date_to_pts()` function is implemented more generally when the package is used for other purposes. When the number of synthetic observations exceeds the number of available observed dates, dates are sampled with replacement from the corresponding empirical temporal distribution.

#### Sampling Through USPopulationSampler Package

**Figure 2.**
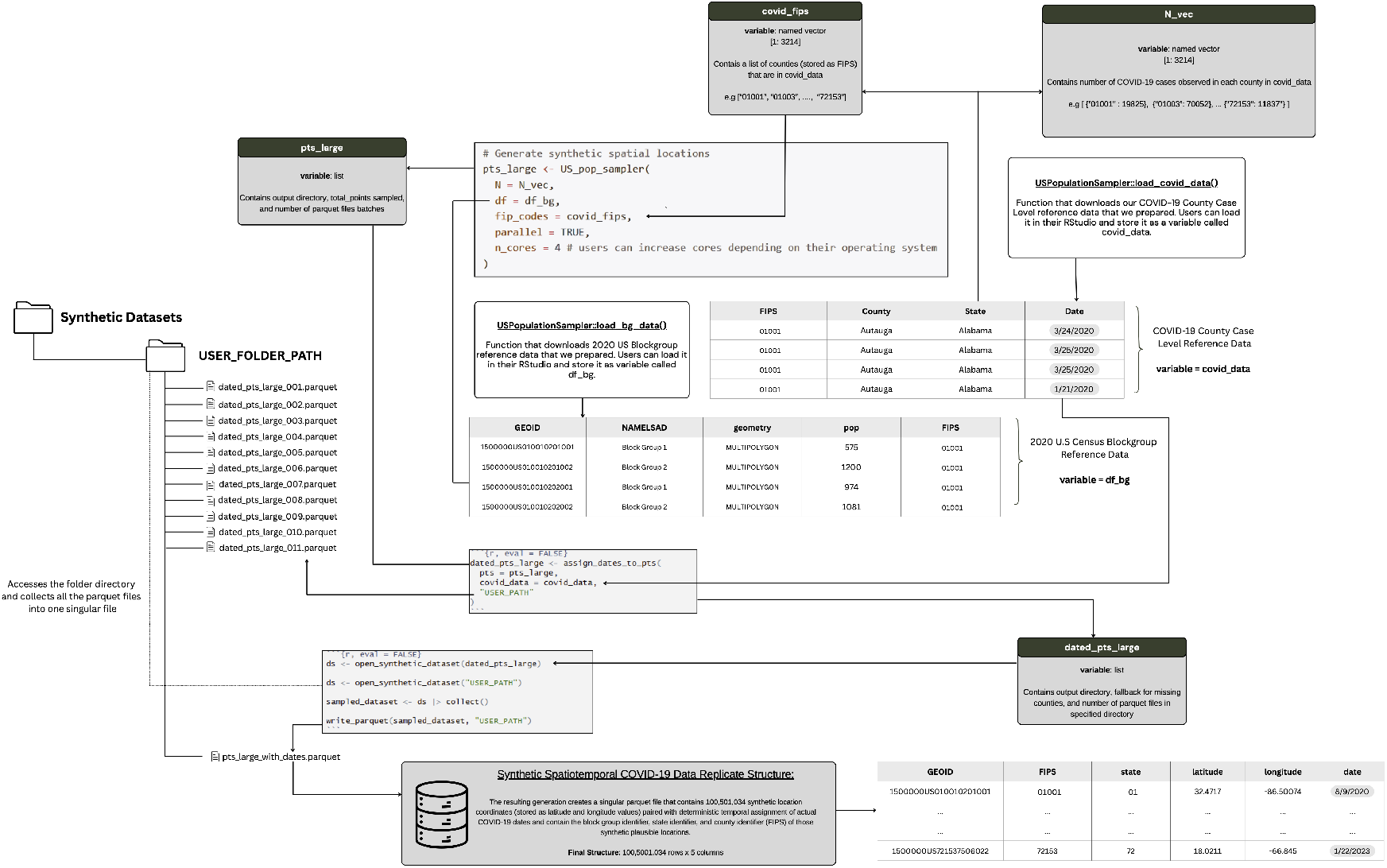
Complete workflow for generating synthetic spatiotemporal Covid-19 replicates using `USPopulationSampler` functions and reference data

~~~
*# Loading the package and necessary libraries*
library(USPopulationSampler)
library(arrow)
library(dyplr)
*# Loading the reference datasets from Zenodo using package’s built-in functions*
covid_data <- load_covid_data(overwrite = **TRUE**)
df_bg <- load_bg_data(overwrite = **TRUE**)
~~~

Users call the library function to load the `USPopulationSampler` package and other necessary libraries into their `Rworkspace` Interactive Development Environment (IDE). Users then load the reference datasets using load_bg_data() and load_covid_data() respectively^24^.

~~~
*# Count observed Covid-19 counts for each county FIPS*
fips_counts <- table(covid_data$fips)
*# Convert to named vector for FIPS-based allocation*
N_vec <- as.integer(fips_counts)
names(N_vec) <- names(fips_counts)
*# Counties observed in the Covid-19 dataset*
covid_fips <- names(fips_counts)
~~~

To reconstruct the synthetic spatiotemporal Covid-19 data replicates, users must first identify all the counties that are present in the Covid-19 reference dataset. The variable `fips_count` stores the number of cases in each fip by using the `table()` function built in base `R`^22^. The variable `N_vec` ensures that the counts are numeric whole integers and the `names()` function ensures that the vector is a named vector. A named vector is a data structure where every element is assigned a label. The variable `covid_fips` is also a named vector which contains the names of all the counties in the Covid-19 reference dataset. Whereas, `N_vec` matches each named county to their numeric number of cases. This avoids the issue of positional matching which can potentially misallocate cases to the wrong county. The `covid_fips` variable named vector should contain 3,214 different counties with 3,210 of those counties existing in the 2020 Decennial U.S. Census Bureau reference dataset^20^. The 4 counties that do not exist are Alaskan placeholder counties `02297` and `02298`, Alaskan discontinued county `02261` for the Valdez-Cordova area, and the Texas missing placeholder county `48999`. These counties and their counts will be reconciliated into other counties within Alaska and Texas.

~~~
*# Generating the synthetic spatial points corresponding to each county*
synthetic_data <- US_pop_sampler(
N = N_vec,
df = df_bg,
fipcodes = covid_fips,
parallel = **TRUE**,
n_cores = 4
)
~~~

The `N` parameter is set to `N_vec` which implicitly sums all the cases present across the counties that will be sampled. This equates to 100,501,034 total sampled points. The data frame parameter takes in the 2020 Decennial U.S. Blockgroup reference dataset and the `fipcodes` parameters matches all the counties FIPS codes present in the Covid-19 reference dataset^20^. The `parallel` parameter activates parallel processing and the number of GPU cores used is 4 to divide the computation into 4 workers to speed up computation time. Depending on the user’s access to high-computing, the number of workers can be increased. Optionally users can specify a numeric seed for reproducibility. Since the number of requested sampled points exceed the streaming threshold of 1 million points, the generated sampled points will be stored as `parquet` files directly into disk to avoid memory explosion in the `Rworkspace` environment. When streaming is triggered in the `US_pop_sampler()` function, the generated sampled data named `synthetic_data` is returned as a list that contains the directory of where the `parquet` files are located, the number of batches written, and the total number of points sampled. Once the synthetic_data has been generated, users can assign dates to the sampled location points using the `assign_dates_to_pts()` function^24^.

~~~
*# Assigning dates to synthetic spatial points*
synthetic_data_dated <- assign_dates_to_pts(
   pts = synthetic_data,
   covid_data = covid_data,
   “USER_PATH” *# replace with own’s directory*
   )
*# Accessing the generated synthetic dataset lazily using Arrow*
ds <- open_synthetic_dataset(synthetic_data_dated)
*# Creating the synthetic data as a single parquet File*
sampled_dataset <- ds |> collect()
~~~

The sampled data generated is passed into the `pts` parameter. The Covid-19 reference data is used for the deterministic assignment of the dates. A directory relative to the user’s personal computer is provided so that dated sampled points are stored as `parquet` files at that location. Users can then use the `open_synthetic_dataset()` function to call on the `arrow` library to lazily access the partitioned `parquet` files as `arrow` objects^27^. Users can employ libraries `arrow` and `dbplyr` functionalities to work with the data lazily without loading into memory^27,28^. If users want all the points in one singular `parquet` file, then the `collect()` function of the `dbplyr` library can be used which combines all partitioned files into one^28^. The following process is repeated to generate the remaining 27 synthetic spatiotemporal Covid-19 datasets that are uploaded onto the specified Zenodo repository^29^.

### Data Record

The 28 synthetic spatiotemporal Covid-19 data replicates are publicly available on Zenodo as `parquet` files via https://zenodo.org/records/21442375 each with a size of 1.8 GB^29^. The datasets can also be accessed via code through

`USPopulationSampler R` package using the `load_synthetic_covid()`^24^.

Each synthetic dataset contains 100,501,034 synthetic individual-level case events of Covid-19 and 5 variable columns:

- `longitude`: longitude coordinate of the synthetic case location, expressed in decimal degrees.
- `latitude`: latitude coordinate of the synthetic case location, expressed in decimal degrees.
- `FIPS`: five-digit Federal Information Processing Series (FIPS) code identifying the county associated with the synthetic case location.
- `GEOID`: census block group identifier corresponding to the block group from which the synthetic location was sampled stored as a 12-digit character string in the format `1500000USsscccTTTTTTB` where `1500000US` is the standard U.S. Census Bureau affiliated geographic identifier, `ss` is the state FIPS code (2-digit), `ccc` is the county FIPS code (3-digit), `TTTTTT` is the census tract code (6-digit), and `B` is the block group number.
- `date`: date associated with the synthetic Covid-19 case event stored as a character string in the format `MM-DD-YYYY`.

The `FIPS` and `GEOID` variables provide geographic identifiers that allow synthetic locations to be associated with their corresponding counties and census block groups. Further descriptions of the data-generation process and reference datasets are provided in the Zenodo repository^29^.

### Technical Validation

The synthetic spatiotemporal data replicates preserve the total number of cases of Covid-19 recorded from 2020 to 2023 across the United States of America. Each dataset contains 100,501,034 total cases which is equal to the number of cases in the NYT Covid-19 dataset spread across 3,210 counties^12^. The original NYT Covid-19 dataset contained 3,214 counties but the extra 4 counties were either placeholder counties or discontinued counties. The synthetic data replicates preserve the territories sampled such as the 50 states, the District of Columbia, and Puerto Rico. The synthetic datasets are further evaluated to assess the preservation of the temporal, geographic, and population distributions observed in the reference Covid-19 dataset. Validation was performed independently for each of the 28 synthetic spatiotemporal data replicates.

Temporal fidelity was determined by observing the daily counts from each synthetic dataset and compared them to the reference COVID-19 dataset. Across all replicates, the mean Pearson correlation coefficient was 0.9999973 while the Spearman correlation coefficient was 0.9999829 which indicates near perfect agreement in both the magnitude and ordering of cases. The mean absolute error (MAE) was 200.257 cases and the mean root squared error (RMSE) was 295.567 cases. This suggests that the deterministic date assignment procedure preserved the temporal distribution of Covid-19 cases with differences occurring due to reconciliated cases receiving dates with replacement when assigned to a new county.

Geographic fidelity was evaluated by comparing county-level case counts between the synthetic and reference COVID-19 dataset. The mean Pearson and Spearman correlation coefficients were 0.99999968 and 0.999998378 respectively which indicates that the synthetic data replicates maintain the geographic distribution of reported cases across U.S. counties with near perfect agreement. The corresponding RMSE and MAE were 86.515 cases and 3.756 cases which indicates minimal differences introduced during the sampling reconciliation procedure. The reconciliation procedure means that there is shifting of some temporal values and exact counts of counties that received reconciliated cases.

The population-weighted sampling algorithm was assessed by comparing the observed proportions of sampled points assigned to each Census block group with the expected proportion derived from the 2020 census population. It appears that the proportion of block groups in the synthetic data replicates and the 2020 Decennial block group reference data matches each other with a Pearson correlation of 0.878 and a Spearman Correlation of 0.872. The corresponding root mean squared error (RMSE) and mean absolute error (MAE) were 1.130 *×* 10^*−*6^ and 6.900 *×* 10^*−*7^ respectively indicating negligible differences between the observed and expected block-group-level proportions.

Overall, the synthetic data replicates maintain high temporal fidelity and geographic fidelity when compared with the COVID-19 data used to construct them. The population-weighted sampling procedure also closely reproduces the expected distribution of sampled locations according to the 2020 Decennial census block group populations. Together, these results demonstrate that the synthetic data generation procedure preserves the principal temporal and geographic characteristics of the Covid-19 reference dataset while simultaneously generating spatially explicit event locations at the census block group level.

**Table 1.** Validation metrics and scope for synthetic spatiotemporal Covid-19 data replicates.

| Validation Component | Metric | Mean ± SD |
| --- | --- | --- |
| Temporal Distribution | Pearson's Correlation | $0.999 \pm 1.332 \times 10^{-7}$ |
| | Spearman Correlation | $0.999 \pm 1.041 \times 10^{-6}$ |
| | Root Mean Square Error (RMSE) | $295.567 \pm 7.354$ |
| | Mean Square Error (MAE) | $200.257 \pm 3.931$ |
| County Distribution | Pearson Correlation | $0.999 \pm 2.093 \times 10^{-9}$ |
| | Spearman Correlation | $0.999 \pm 4.372 \times 10^{-8}$ |
| | Root Mean Square Error (RMSE) | $86.515 \pm 0.281$ |
| | Mean Square Error (MAE) | $3.756 \pm 0.000$ |
| Population-Weighted Sampling | Pearson Correlation | $0.878 \pm 6.270 \times 10^{-5}$ |
| | Spearman Correlation | $0.872 \pm 6.170 \times 10^{-5}$ |
| | Root Mean Square Error (RMSE) | $1.130 \times 10^{-6} \pm 2.610 \times 10^{-10}$ |
| | Mean Square Error (MAE) | $6.900 \times 10^{-7} \pm 1.190 \times 10^{-10}$ |

## Data Availability

All data produced in the present study are available upon reasonable request to the authors.

https://zenodo.org/records/21442375

## 1 Data Availability

Data can be downloaded through the Zenodo repository page https://doi.org/10.5281/zenodo.21442375 ^29^. The repository contains two reference datasets that the package is built upon and 28 synthetic spatiotemporal Covid-19 data replicates. The first reference dataset is named `US_Census_Blockgroup_Data_2020.rda` which is a cleaned 2020 Decennial Census data titled `Redistricting Data / Public Law 94-171` by extracting block group population and information of counties across the United States of America^20^. The second reference dataset is named `Covid_19_County_Case_Data.rda` which is a cleaned event-level Covid-19 data extracted from the aggregated county level Covid-19 data published by the NYT^12^. Lastly, the data replicates are called synthetic_data with a number at the end indicating their numerical listing order which includes synthetic_data_1 up to synthetic_data_28.

## 2 Code Availability

All code is hosted freely and open-source on a GitHub repository called https://github.com/Techavoan/USPopulationSampler. The software version used is `R` version 4.5.2 (2025-10-31) and the release version of `USPopulationSampler` is 1.0.0 undergoing CRAN submission.

## Acknowledgments

AJH is supported by NSF grant DMS 2236854 and NIH grant R35 GM159431.

## Author contributions statement

RY simulated the datasets, wrote the manuscript, and developed the entire package. AJH produced the concept, provided oversight on the whole project. JZ helped provide mathematical insights and worked on a prototype version of `USPopulationSampler` in Summer 2025. All authors reviewed the manuscript.

## Competing interest statement

The author(s) declare no competing interests.

## Notes

### Competing Interest Statement

The authors have declared no competing interest.

